# The DiaGame Study: Multimodal Data Collection in Free-Living Conditions Using Consumer-Level Wearable Devices and Apps in People with Diabetes

**DOI:** 10.1101/2024.12.13.24318201

**Authors:** Ryan de Vries, Edouard F.H. Reinders, Danilo Ferreira de Carvalho, Elma M.W.L. Cruts – Van de Meulengraaf, Pieter M.E. van Gorp, Pleun C.M. Wouters - van Poppel, Uzay Kaymak, Peter A.J. Hilbers, Natal A.W. van Riel, Harm R. Haak

## Abstract

The everlasting interest in precision medicine has instigated a need for patient-specific multidimensional data collection. By leveraging consumer-level wearable devices and applications we can support this need by collecting multimodal data in uncontrolled free-living conditions. We used a continuous glucose monitor, a smartwatch with in-house developed application, and a smartphone application over a period of 14 days to collect data from 60 participants with diabetes. Data were collected on 77,683 CGM measurements, 4,073 moods, 3,517 meals, 1,394 insulin injections, and 1,364 bouts of physical activity. Participants were able to effectively use the devices and applications, with 45 participants reporting data on all modalities and all participants reporting data on at least two modalities (in addition to the data collected from the smartwatch’s internal sensors). These findings highlight the efficiency and effectiveness of consumer-level wearable devices to collect data from the comfort of the people’s homes. Thereby, facilitating the need for a cost-effective and scalable approach for precision monitoring to the public.

## Introduction

The emergence of precision or personalized medicine, which seeks to improve diagnosis, prediction, prevention, and treatment by focusing on the individual rather than on groups of people, has instigated a need for patient-specific multidimensional data to evaluate an individual’s health, wellbeing, and environment. Diabetes mellitus, a chronic endocrine disease characterized by chronically elevated blood glucose levels, is one example of a disease for which precision medicine approaches can prove to be beneficial, due to the recognized heterogeneity in its etiology, clinical presentation, and pathogenesis^1^. The resulting need for patient-specific multidimensional data collection (precision monitoring) in people with diabetes has been highlighted in the latest American Diabetes Association (ADA) and European Association for the Study of Diabetes (EASD) consensus, along with recent publications, that advocated for precision monitoring methods as the next crucial step towards precision medicine for diabetes^1,2^. Precision medicine in the field of diabetes research focuses on elucidating pathophysiological processes (molecular and environmental), and consequently the heterogeneity in glycemic dynamics, that underly the disease. Although the primary cause of dysregulated glycemic dynamics concerns defects in at least one physiological pathway, multiple coherent factors including self-regulated behavior (e.g., dietary intake and physical activity) and health (e.g., medication and psychological status) also influence glycemic dynamics. Furthermore, lifestyle, defined by the long-term combination of these variables, impacts the risk for future complications^3–7^.

To capture glycemic variations, gain insight into the underlying glycemic effects of lifestyle, and ultimately monitor disease progression, all at an individual level, the availability of data is imperative. Although data measured in clinical and research settings are well controlled, detailed data collection over weeks or months of time is unfeasible from these settings and difficult to scale to large cohorts. Consequently, a transition must be made towards data measured in the comfort of people’s homes and throughout their daily lives. Furthermore, data from free-living conditions are representative of self-regulated behavior and habits, while measurements in controlled clinical settings are not, and these data offer the opportunity to capture the day-to-day glycemic variability typical for diabetes. The shift from clinical settings towards free-living conditions, to collect data for healthcare and research purposes, is enabled by the advent and enhancement of wearable devices. The continuous glucose monitor (CGM) has already provided tremendous insight into glycemic control for clinicians and people with diabetes^8^. Furthermore, smartphones and smartwatches have been shown to be promising for the monitoring of lifestyle variables (i.e., assessment of physical activity, diet, and psychological stress)^9,10^.

The use of wearable devices in diabetic populations for data collection in free-living conditions has, except for a few studies, been mostly limited to the effects on blood glucose levels of physical activity, diet, or psychological stress alone. The limited number of studies that simultaneously collected data on a broader range of variables associated with glycemic variability, have almost all been conducted with people with type 1 diabetes. Marling & Bunescu (2020) collected data in 12 participants with type 1 diabetes for eight weeks on blood glucose levels (CGM), dietary intake, physical activity, usage of insulin analogues and other antidiabetic medication, and psychological status in free-living conditions^11^. Other studies such as the *D1NAMO project* (nine participants with type 1 diabetes, four-day study period) have collected data on electrocardiograms, breathing, accelerometers, CGMs, and annotated food pictures^12^. The *ShanghaiT1DM* & *ShanghaiT2DM* datasets (12 participants with type 1 diabetes and 100 participants with type 2 diabetes, study period up to 14 days) included dietary intake, insulin injections, and CGM data^13^. Nevertheless, while these studies collected multimodal data in free-living conditions in people with type 1 diabetes, similar studies including people with type 2 diabetes are rare. Zahedani et al. (2023) used a CGM, a proprietary app for food logging, and heart rate data from a smartwatch to demonstrate that app-based lifestyle recommendations can improve eating habits, markers of glycemic variability, and weight in a predominantly healthy and prediabetic cohort (N = 1,066, including 94 subjects with non-insulin-treated type 2 diabetes)^14^. Similarly, Pai et al. (2024) collected data from a CGM, app-based food logging, and accelerometer-derived step count to stratify postprandial peaks into normal and elevated categories and highlighted the relationship between post-meal steps and a reduced postprandial glucose response in Hispanic/Latino adults (N = 36, including 14 subjects with non-insulin-treated type 2 diabetes)^15^.

Data collection in free-living conditions can be burdensome for the participants and researchers, especially when using analog data collection methods (i.e., paper logbooks and questionnaires). The data collection process often requires data digitization, structuring, and manual preprocessing. Some studies also relied on participants or experts to acquire information on dietary macronutrient composition. Dubosson, et al. (2018) used picture-based food annotation by a dietitian, while Marling & Bunescu (2020) relied on patient estimates to infer carbohydrate intake.

By integrating multiple wearable devices, and applications, some of the burdens of data collection in free-living conditions can be alleviated. The use of wearable devices negates the need for data inference and digitization. Moreover, the use of specifically consumer-level wearable devices, rather than research-dedicated devices, offers a cost-effective, scalable, and readily available method of collecting data due to the widespread ownership of smartphones and the increasing popularity of smartwatches and fitness trackers. The consumer-level wearable devices, used in this study, included a CGM for continuous blood glucose measurements, a smartphone application to collect patient-reported dietary intake and physical activity, and a smartwatch to measure heart rate, acceleration, and step counts. The smartwatch is supplemented with an in-house developed application (publicly available) to allow participants to self-report mood and insulin usage. This application also incorporates a nudging mechanism, using vibrations and visual prompts on the screen, to encourage participants to report information as requested by the research team. The use of consumer-level wearable devices ultimately enables large-scale, patient-centered, and researcher-independent personalized data collection.

This article describes how commercially available consumer-level wearable devices and apps were used to collect data in free-living conditions in people with diabetes, predominantly type 2 diabetes. Subsequently, we assessed the feasibility of using wearable devices in this study population, which is typically older, and analyzed the effectiveness of our approach in terms of both the quantity and quality of the data collected. These data included measurements of blood glucose levels, detailed dietary intake, physical activity, insulin use, and mood over a 14-day period, supplemented with demographics and clinical measurements. Ultimately, we aim to showcase the ease and extent to which multimodal data can be collected in free-living conditions using consumer-level wearable devices and applications in a diabetic population (predominantly type 2).

## Results

### Participant Characteristics

Upon study completion, 59 participants (35 males and 24 females) were included (excluding one dropout due to multiple CGM malfunctions). For one participant CGM data of her own prescribed CGM was voluntarily provided due to device malfunction. 51 participants were diagnosed with type 2 diabetes and eight with type 1 diabetes, with respective mean ages of 69·1 years (SD 7·1) and 54·6 years (SD 16·9), mean BMIs of 30·1 kg/m^2^ (SD 4·9) and 25·4 kg/m^2^ (SD 3·3), mean diabetes durations of 20·5 years (SD 7·3) and 26·6 years (SD 13·4), mean HbA1c’s of 61·5 mmol/mol (SD 11·7) and 63·4 mmol/mol (SD 15·0), and mean time in ranges (3·9-10·0 mmol/L) during the study of 66·8% (SD 22·7) and 62·7% (SD 21·6). For a complete overview of participant characteristics (e.g., insulin and medications use, anthropometric measurements, and fasting blood values), stratified by diabetes type, see Table 1.

**Table 1:** Participant characteristics stratified by diabetes type. *n=58 as one participant had not fasted the morning of visit 2. BLOQ: Below Limit of Quantification (0·03 nmol/L for C-peptide); these values were imputed with half of the detection limit concentration to calculate the corresponding presented statistic. Time in range is defined by the percentage of times CGM values were in between 3·9 and 10·0 mmol/L.

| Demographics | Mean ± Standard Deviation |  |
| --- | --- | --- |
|  | Type 2 Diabetes<br>(n = 51) | Type 1 Diabetes<br>(n = 8) |
| General |  |  |
| Male | 32 | 3 |
| Female | 19 | 5 |
| BMI (kg/m<sup>2</sup>) | 30.1 ± 4.9 | 25.4 ± 3.3 |
| Age (years) | 69.1 ± 7.1 | 54.6 ± 16.9 |
| HbA1c (mmol/mol) | 61.5 ± 11.7 | 63.4 ± 15.0 |
| HbA1c (%) | 7.8 ± 1.1 | 8.0 ± 1.4 |
| Systolic Blood Pressure (mmHg) | 133 ± 16 | 128 ± 13 |
| Diastolic Blood Pressure (mmHg) | 73 ± 8 | 76 ± 4 |
| Resting Heart rate (bpm) | 75 ± 17 | 81 ± 12 |
| Diabetes Duration (years) | 20.5 ± 7.3 | 26.6 ± 13.4 |
| Fasting Glucose (mmol/L) | 7.8 ± 2.5* | 10.7 ± 4.0 |
| Fasting C-Peptide (nmol/L) | 0.69 ± 0.44*<br>(1 BLOQ) | 0.10 ± 0.14<br>(5 BLOQ) |
| Time in Range During the Study (%) | 66.8 ± 22.7 | 62.7 ± 21.6 |
| Daily Insulin per kg bodyweight<br>(IU/kg/day) | 0.45 ± 0.45 | 0.44 ± 0.20 |
| Insulin Analogues and Medication |  |  |
| Insulin Aspart | 17 | 7 |
| Insulin Lispro | 2 | 1 |
| NPH Insulin | 2 | 1 |
| Insulin Glargine | 9 | 0 |
| Insulin Detemir | 2 | 0 |
| Insulin Degludec | 25 | 5 |
| Mixed Insulins | 1 | 0 |
| Insulin Pump Therapy | 1 | 2 |
| Metformin | 41 | 1 |
| SGLT2 Inhibitors | 17 | 0 |
| GLP-1 Agonist | 23 | 0 |
| Sulfonylureum Derivatives | 25 | 0 |

### Data Quantity

The set of wearable devices provided a large extent of multimodal data (Fig. 1 showcases an example of four days of collected data). Data in Fig. 1 includes sensor measurements of interstitial glucose concentrations and heart rate as well as self-reported mood, insulin analogue injections, dietary intake (only carbohydrate content is shown here), and physical activity. The data not only provided insight into the day-to-day glucose variability at an individual level, but glycemic variations can also be associated with reported self-regulated behavior. In this example, rises in blood glucose levels were associated with high-carbohydrate meals and drops with adequate preprandial insulin administration or periods of reported physical activity with an elevated heart rate. In total, the dataset includes 77,683 CGM measurements, 4,073 self-reported moods, 3,517 meals, 1,394 insulin injections, and 1,364 bouts of physical activity. Excluding days of study visits, interstitial glucose levels were measured (every 15 minutes) in 97·5% of measurement instances. A daily energy intake of at least 1,000 kcal/day was reported on 80·6% of study days (excluding a single participant who was known to have started a crash-diet prior to study participation), while at least 1,000 steps a day were recorded on 93·0% of study days (excluding one wheelchair-bound participant). 96·6% of participants self-reported their mood, and 84·1% of insulin-dependent participants (n=44) reported insulin doses. 84·5% of participants had self-reported physical activity. 45 participants (76·3%) managed to self-report data of every modality (including insulin doses when prescribed). Further analysis demonstrated participants were consistently engaged (i.e. they reported data) with the applications and smartwatch during the study period, only with a slight decrease in physical activity reporting after the second and third study days (Supplemental Fig. S3). The number of days participants reported data on each modality varied between participants (shown in Fig. 2). Excluding visit days, dietary intake had been reported for 11 to 13 days by all participants (67·8% with a daily energy intake greater than 1,000 kcal/day). The number of days with bouts of physical activity, which had a qualitatively similar distribution to the number of days with at least 3,000 steps, ranged from between nine and 13 days (total of 54·2%) to only one or no day (total of 27·1%). Mood was self-reported anywhere between zero (3·4%) and 13 (22·0%) days, with more than half (50·8%) of the participants reporting mood for at least ten days. 70·5% of insulin-dependent participants reported insulin usage for 11 to 13 days, while seven out of 44 (15·9%) of these participants (only one of these seven used rapid-acting insulin) did not report insulin usage at all.

**Figure 1:**
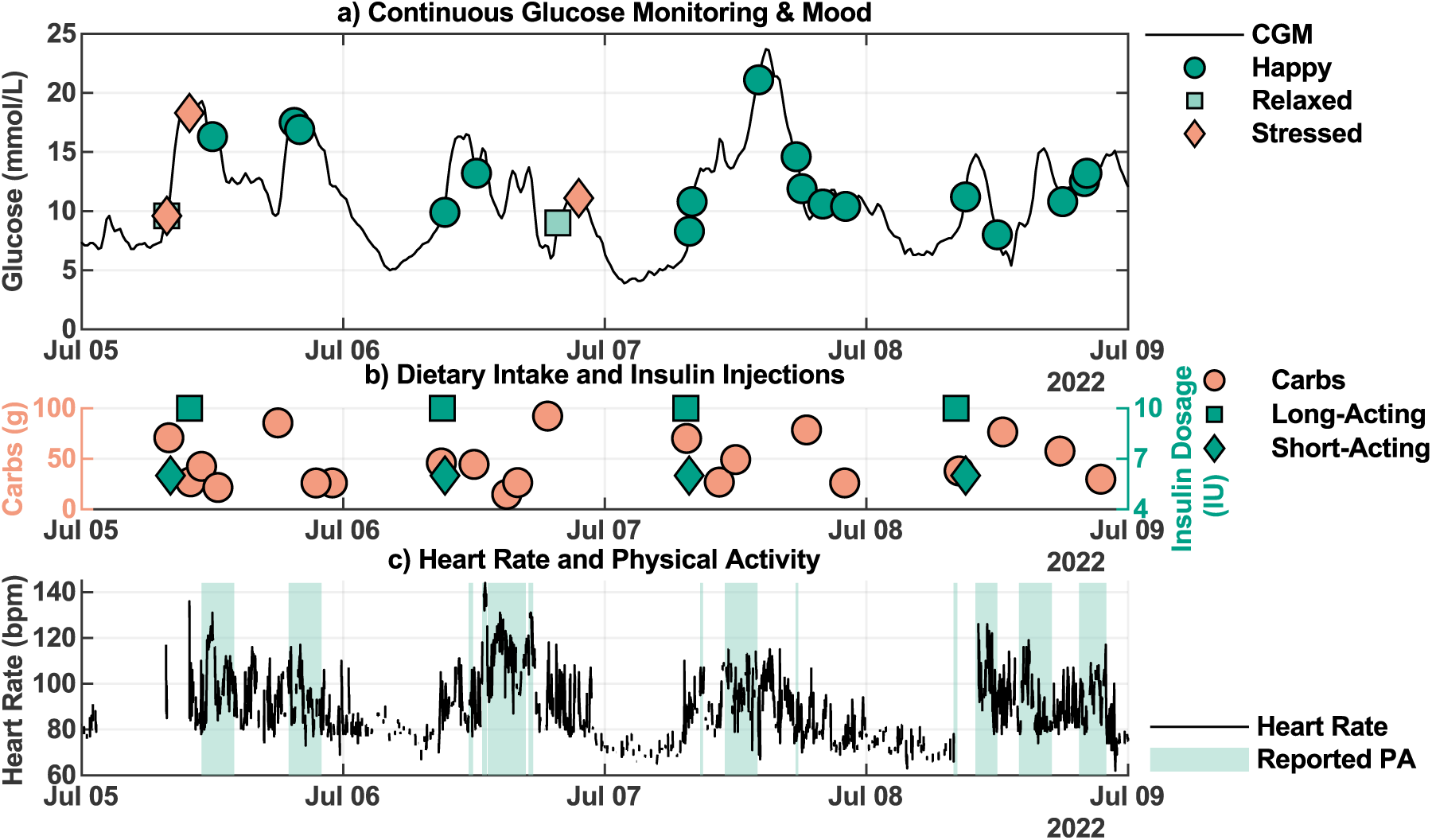
Example of four days of data collected from one participant. Panel A: CGM measurements (in mmol/L, solid black line) and reported moods (dark green circle = happy, light green square = relaxed, light red diamond = stressed; sad and angry were not reported by this participant in this interval). Panel B: Carbohydrate content (in grams) of reported dietary intake (light red circles) and reported short-acting (dark green diamonds) and long-acting (dark green circles) insulin doses (IU). Panel C: Measured heart rate in beats per minute (solid black line) and participant-reported bouts of physical activity (light green shade). Accelerometer and pedometer data were excluded from this figure for clarity.

**Figure 2:**
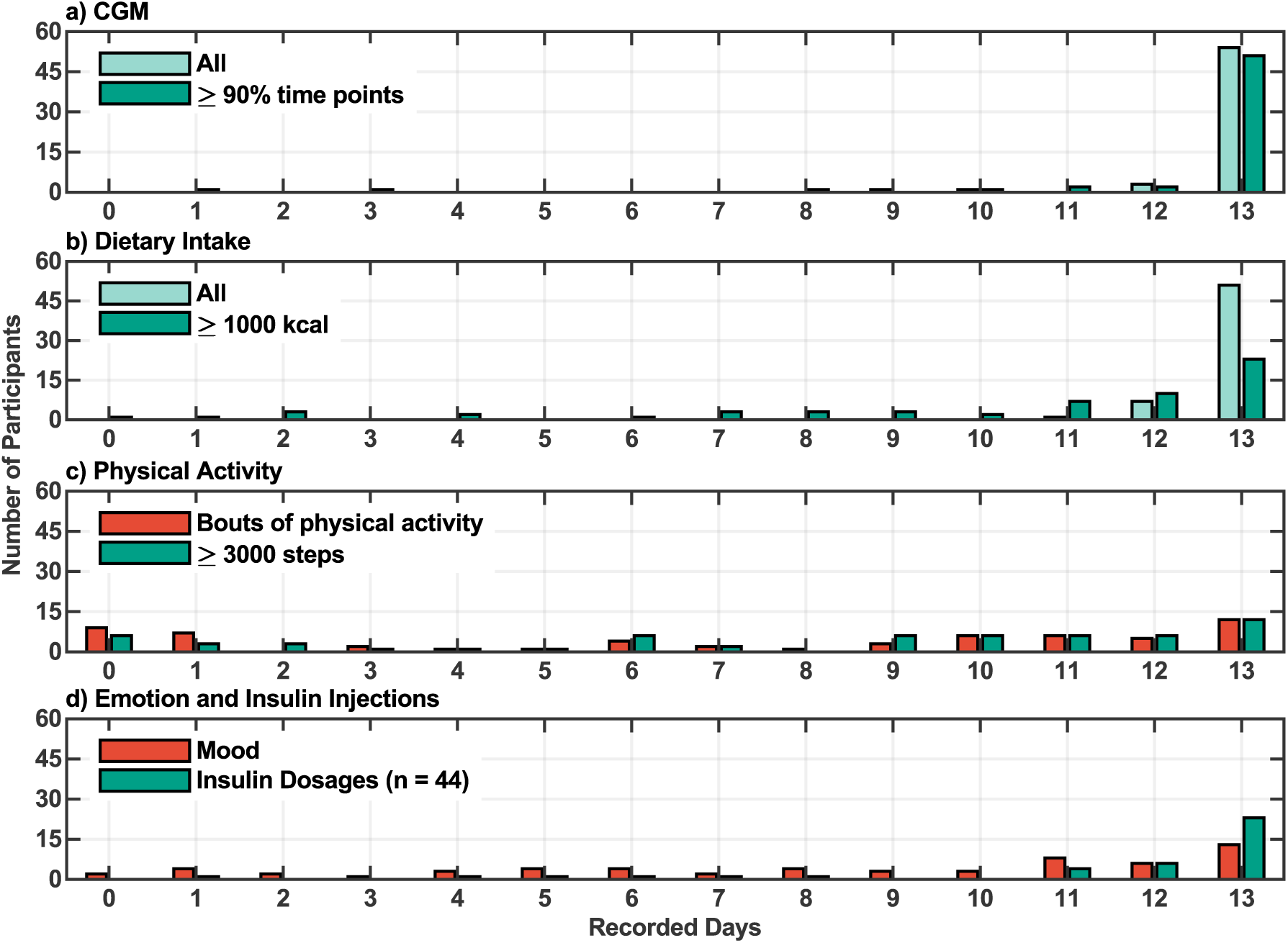
Distribution of available days of collected data per participant for various data modalities. Distribution of the total number of days, excluding days of study visits, with collected data per participant for each of the data modalities. Panel A: Distributions of available CGM measurements (including all days = light green, including days with at least 90% of CGM time points = dark green). Panel B: Distributions of self-reported dietary intake (including all days = light green, including days with at least 1,000 kcal reported = dark green). Panel C: Distributions of self-reported physical activity (red) and days with at least 3,000 steps (green). Panel D: Distributions of self-reported mood (red) and insulin injection doses (green, n = 44 since not all study participants were insulin dependent).

### Data Quality

Evaluation of the quality of the patient-reported data suggested mixed results, as data was consistently reported but not always to the extent that it matched our expectations of a complete day of lifestyle events. A comparison of reported dietary intake and daily insulin doses to recommended guidelines and health records demonstrated discrepancies for a subset of participants. Firstly, the reported daily energy intake (shown in Fig. 3, with participants ordered by age), varied considerably between participants from 55·2 kcal/day to 4,766 kcal/day. The mean energy intake (excluding outliers below 1,000 kcal/day) was 1,796 kcal/day (SD 563 kcal/day) for men (dashed blue line) and 1,484 kcal/day (SD 356 kcal/day) for women (dashed green line). For reference, dietary reference values recommend 1,860 and 2,290 kcal/day for, respectively, moderately active females and males between the ages of 60 and 69 years^16^. Thus, the reported energy intake was lower than these reference values. A complete breakdown of macronutrient intake (carbohydrates, sugars, saturated and unsaturated fat, protein, fiber, and salt) is provided in Supplemental Fig. S4. Additionally, mean reported daily energy intake was not correlated with year of birth (Spearman’s ρ = -0·19, p = 0·15).

**Figure 3:**
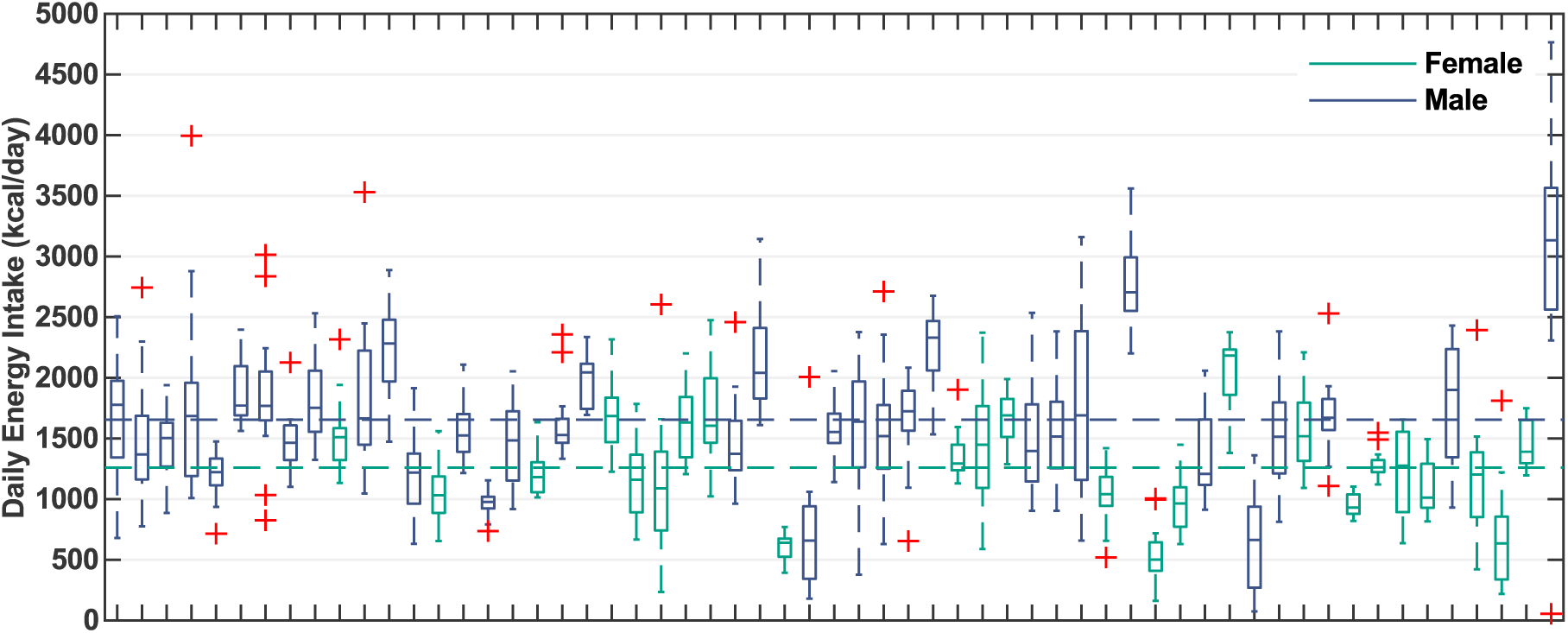
Reported daily energy intake colored by sex and sorted by year of birth (ascending from left to right). Boxplots of the daily energy intake (kcal/day) reported from day two (day one excluded due to the variability of the times of day at inclusion) until visit 2 for each participant. Participants are colored by sex (blue = male, green = female, outliers are colored red) and sorted by year of birth (ascending from left to right). Mean daily energy intake per sex (including outliers) is depicted by corresponding dashed lines (1,655 kcal/day for males and 1,259 kcal/day for females).

Secondly, Bland-Altman plots comparing daily (accumulated) reported insulin doses with prescriptions from health records, stratified by rapid and (ultra) long-acting insulin analogues (Supplemental Fig. S5 and S6), showed an overall mixed agreement between reported and prescribed values. The median bias of respectively rapid- and long-acting analogue doses were both 0 IU/day. The relative difference between reported and prescribed daily dose was between -15% and 15% for 50·2% of study days for short-acting insulins (this range was defined to allow some daily variability, typical for short-acting insulin treatment). There was no difference between reported and prescribed dose on 55·5% of study days for long-acting insulins. Otherwise, daily insulin doses were mostly underreported compared to prescribed doses (respectively 31·6% and 31·0%). Note for example the distinct declining line of points for long-acting insulins that correspond to days without reported doses. Nonetheless, the deviations in the reported daily doses could be attributed to a subset of individuals (Supplemental Fig. S7 and S8).

Ecological momentary assessment of mood with the nudging mechanism using the smartwatch provided a method to repeatedly gain insight into psychological status within the natural flow of life. Of the five mood categories, participants primarily reported being happy (13·5%) or relaxed (78·9%) throughout the day (Fig. 4), while stressed (5·4%), sad (1·2%), and angry (1·0%) were reported less frequently. Moreover, mood was reported most frequently shortly after participants were nudged, evident by a considerably higher number of reports during the hours of the day including 08:00, 12:00, 18:00, and 20:00. Differences in the number of reports at these four times can be attributed to differences in wearing percentage and response percentage. Firstly, the wearing percentage was lower at 52·7% (95% CI, 49·1% – 56·3%) in the morning compared to 94·0% (95% CI, 92·1% – 95·6%), 95·6% (95% CI, 93·9% – 96·9%), and 96·1% (95% CI, 94·5% – 97·3%) at 12:00, 18:00, and 20:00, respectively. Secondly, the nudging response percentage was considerably lower at noon with 33·0% (95% CI, 29·6% – 36·6%) compared to 18:00 and 20:00 with respectively 41·5% (95% CI, 37·9% – 45·1%) and 42·1% (95% CI, 38·5% – 45·7%), while the response percentage at 08:00 was 35·4% (95% CI, 30·7% – 40·3%). Thus, the nudging mechanism was most effective in the evening times. These wearing and response percentages, overall and stratified by nudge time, did not decrease throughout the study period and were unaffected by the check-up call around study day five (Supplemental Fig. S9 and S10). Nevertheless, similar to the reported dietary intake and insulin doses, the response percentages varied considerably among participants, ranging from 0% to 96% during the observational period (Supplemental Fig. S11). However, the percentages of responses to the nudging were not correlated with the number of moods reported outside of the nudging times (Spearman’s ρ = -0·08, p = 0·53).

**Figure 4:**
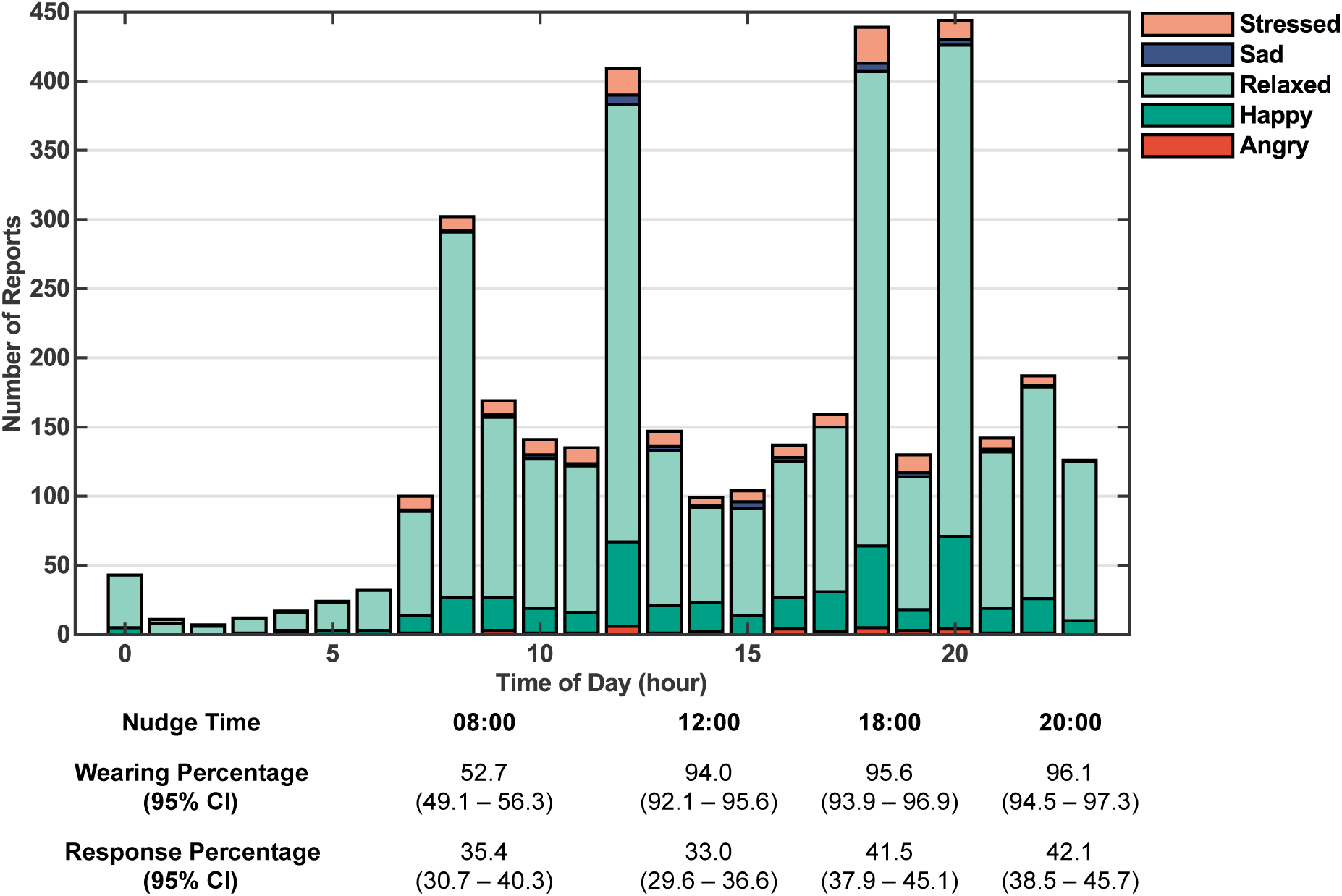
Moods reported by participants at each time of day and nudging statistics at each nudging instance. Number of moods reported at each hour of the day (angry = dark red, happy = dark green, relaxed = light green, sad = dark blue, stressed = light red). Underneath the figure, the wearing percentage at each nudge time (08:00, 12:00, 18:00, and 20:00) and corresponding response percentage to the nudges are given with 95% confidence intervals.

## Discussion

By leveraging consumer-level wearable devices, supplemented with applications and a nudging mechanism, we aimed to efficiently collect multimodal personal data in free-living conditions on glycemic dynamics and an extensive set of associated variables (dietary intake, insulin administration, physical activity, and mood). Furthermore, the effectiveness and feasibility of using multiple wearable devices (CGM, smartphone, and smartwatch) with corresponding applications was assessed for data collection in a study population typical of type 2 diabetes (relatively old age and various comorbidities).

Our study showcased the opportunity to harness wearable devices as a feasible method of multimodal longitudinal data collection under free-living conditions in individuals. These devices required minimal calibration steps, could be operated by participants following a short instruction session, and no major problems were identified by the research team (except for two CGM sensor failures) or reported by participants. Moreover, the interaction of the participants with the applications was effective, reflected by the high percentage of participants providing self-reported information. Additionally, the use of a blinded CGM negated the need to scan every eight hours, potentially leading to loss of data and avoided influencing the participant’s decision-making. The commercial availability of these consumer-level devices and public access to the applications allows for time-effectiveness and easy scalability to large cohorts. These devices, thereby, provide the means to obtain continuous insight into lifestyle and personalized glycemic dynamics to the public and optionally their healthcare providers.

The combination of a CGM, a smartwatch, and a smartphone application provided insight into the daily life of an individual, from the comfort of their home, of which limited information was otherwise available. However, data collected under free-living conditions is usually not a comprehensive description of all lifestyle events. Evaluation of data quality indeed indicated that participant-reported data did not always compare to expectations, with heterogeneity in the magnitude of these differences between participants. Firstly, the energy intake reported through the smartphone application (Fig. 2) was, on average, lower than expected (around 450 kcal/day lower) considering the European guidelines for moderately active women and men between the ages of 60 and 69 years^16^. Nonetheless, the mean reported energy intake was comparable to another study using the same smartphone application in a younger non-diabetic population (1,830 kcal/day, SD 485 kcal/day, n=100), which concluded that the application provided comparable results to 24-hour recalls. Dubosson et al., whom relied on picture-based food annotation by a dietitian, reported similar daily energy intakes (1,672 kcal/day, SD 1,038 kcal/day, n=8)^12,17^. Additionally, mean daily carbohydrate intake was similar to participant-reported values in people with type 1 diabetes^11^. Therefore, even though the energy intake may deliberately be reduced or underreported by participants during a study focused on lifestyle observation, partially accounting for the discrepancy, accurate dietary intake assessment in free-living conditions remains challenging. Nonetheless, all participants had reported dietary intake for at least 11 days. The participant’s age did not appear to be a limiting factor in reporting dietary intake using a smartphone application, even with a generally older cohort typical for type 2 diabetes.

Secondly, the smartwatch offered an easily accessible means for insulin usage registration, which needed no connection to the internet or a smartphone and was always at hand for participants. These reported insulin doses were accurate on a majority of the study days, when they could be compared to prescribed insulin treatments. These comparisons demonstrated that only five out of 36 long-acting insulin users, and no short-acting insulin users, failed to report at least one day for which the accumulated daily insulin dosages were comparable to their health records. However, the accumulated daily insulin dosages derived from insulin reports were frequently lower than prescribed (especially evident when one dose a day was prescribed, and none were reported). It is unclear why insulin administration was underreported. While daily dose adjustments are inherently part of the short-acting insulin treatment, the quality of reported data on insulin use may be affected by the otherwise repeated nature of daily insulin dosing leading to a lack of urgency to consistently report these events. It has already been shown that data on insulin dosing can be effectively collected from insulin pumps^18^, which are predominantly used by people with type 1 diabetes, whereas the recent development of smart insulin pens could potentially alleviate the burden of insulin dose registration altogether.

Thirdly, ecological momentary assessment of mood using the smartwatch, supplemented with a nudging mechanism four times a day, appeared to be an effective method to collect information on the perceived mood of participants. Participant involvement was consistent throughout the study period with high wearing percentages (except in the early morning when several participants may not yet have woken up) and overall steady response percentages. Participants for whom the nudging mechanism was less effective still provided ample mood reports without this nudging encouragement. In addition to providing a means to report mood and insulin doses, the smartwatch enabled the measurement of heart rate, acceleration, and step count to independently inform on physical activity (next to participant-reported physical activity). Lastly, the mean daily value of none of the variables reported by the participants decreased considerably throughout the study period; which, if this had happened, could have suggested that the data collection protocol may have placed too much of a burden on the participants. Thus, while the collected data may not be a complete description of all daily events, these wearable devices can effectively be used to acquire multimodal data in free-living conditions. Hereby answering the call for precision monitoring methods as the next step towards precision prevention, diagnostics, and prognostics ^1,2^.

We acknowledge that the study cohort consisting of known patients does not provide a complete cross-sectional overview of the diabetic population in the Netherlands. Nonetheless, the study cohort is typical for a type 1 and 2 diabetes cohort, when compared to other studies in terms of patient characteristics. Inclusion may be biased for participants with at least some affinity for digital devices as those unfamiliar or hesitant to use wearable devices may be more likely to refuse participation. These latter people may need additional support to promote adequate use of these wearable devices.

Multiple enhancements could further enrich the collected data through the inclusion of standardized at-home meals and meal tolerance tests^19^. Furthermore, the registration of medication usage can be extended from the use of solely exogenous insulin to oral medications (e.g., metformin and GLP-1 agonists). Adherence to these medications was currently assumed to be consistent throughout the study period, even though in practice adherence has shown to be inconsistent^20^. Additionally, real-time communication between devices can enable personalized nudging mechanisms, by incorporating information from all devices, for Just-In-Time Adaptive Interventions^21^. Tailoring the timing, frequency, and format of the nudging to the individual may improve effectiveness, while just-in-time nudging could be used to promote reports on dietary intake or insulin injections when blood glucose excursions are observed (currently the nudging mechanism was limited to data collection on mood due to the inherent continuity of human mood, which is lacking in all other variables).

The data collected in free-living conditions holds the potential for various clinical purposes, as indicated by the growing interest and recent increase in research within this field. Firstly, it offers a deeper understanding of intra-day and inter-day glycemic variability at an individual level. This understanding enables healthcare providers to deliver targeted and personalized care (precision medicine), which may contribute to improved health outcomes (e.g., to deliver app-based personalized lifestyle recommendations^14^). Secondly and more importantly, the collected data offers valuable insights when analyzed across broader groups. It can reveal inter-individual variations, facilitating the identification of distinct subgroups within the diabetes population, unveiling their distinct characteristics and needs (e.g., to stratify meal events for digital phenotyping^15^). Lastly, these data can support the development of next-generation simulators and digital twins, general or personalized dynamic metabolic models capable of predicting glucose levels and their modulation due to daily activities^22^. Such models have the potential to serve multiple purposes, from enhancing broader group-level education by showcasing the response of blood glucose levels to lifestyle decisions to *in silico* experiments with virtual patient cohorts. A deeper understanding of a disease like diabetes and the various factors influencing blood glucose levels can foster increased self-confidence and effective self-management in people with diabetes. Given that improved self-management in diabetes correlates with better health and reduced short- and long-term complications, the significance of this understanding is clear^23^. Furthermore, diabetes self-management education and support (DSMES) currently occurs predominantly through one-on-one consultations with healthcare providers. However, with the escalating numbers of patients with diabetes globally, there is a growing need for more scalable, sustainable, and affordable support. Electronic resources and digital interventions, such as advanced simulators, predicting glucose levels at a personal level, could offer a crucial addition to future healthcare. They hold the potential to make healthcare more efficient and accessible while simultaneously improving its quality.

## Conclusion

Our study showcased the feasibility and effectiveness of consumer-level wearable devices for multimodal data collection in free-living conditions, in a predominantly type 2 diabetes cohort. Moreover, the protocol can be scaled to larger cohorts and easily reproduced and integrated due to the commercial or free availability of the devices and applications. The multimodal data that can be measured in free-living conditions, by relying on wearable devices, applications, and the people themselves, give us the opportunity for precision monitoring to individualize patient care by accounting for intra- and inter-day heterogeneity in glycemic dynamics. Moreover, the data support further research into the effects of self-regulated behavior and health on glycemic dynamics and the long-term progression of the disease. Ultimately, enabling us to provide patients with personalized approaches for diabetes treatment and self-management support.

## STAR Methods

### Study Design and Participants

Between June 2022 and June 2023, 60 participants (both type 1 and type 2 diabetes) were included and data were collected over a consecutive 14-day period. All participants were recruited from the outpatient clinic of the Máxima Medical Centre (The Netherlands). Participants were eligible for inclusion when they met the following criteria: (I) diagnosis of type 2 diabetes or type 1 diabetes (body mass index (BMI) < 30 kg/m^2^ for type 1 diabetes), (II) age > 18 years, (III) possession of a smartphone that runs the required application. Exclusion criteria comprised: (I) pregnancy or breastfeeding, (II) ongoing treatment for malignancy, (III) scheduled MRI scan during the study, and (IV) non-Dutch speaking. All individuals provided written informed consent. The study was registered in the Dutch Trial Registry (NL9290), approved by the Medical Research Ethics Committee Máxima MC (nr. L20.102), and conducted according to the principles of the Declaration of Helsinki.

Participation consisted of two visits to the clinic and a telephone call. During the first visit, a brief overview of the participant’s medical history was registered, along with medication use. Subsequently, blood pressure, resting heart rate, weight, and length were measured and a blinded CGM was applied. Finally, participants received a smartwatch with preinstalled study application, were instructed on how to download the smartphone application, and use the devices.

Four to six days after the first visit participants were contacted by telephone to address specific issues, identified through a remote assessment of data reported on dietary intake and physical activity. Instructions were reiterated if necessary. Subsequently, after 14 days, participants returned for the second visit in a fasted state to draw blood and return all devices. The data on these devices were subsequently pseudonymized and exported to a secure database.

### Data Collection

Glucose concentrations were measured using a *FreeStyle Libre Pro iQ* sensor (Abbott Diabetes Care, Illinois, USA), a blinded CGM that measured interstitial glucose levels every 15 minutes, 24 hours per day, for 14 consecutive days. Glycated hemoglobin (HbA1c), fasting glucose, and c-peptide concentrations were measured using the standard protocol of the hospital’s clinical chemical laboratory.

Participants received a *Samsung Galaxy Watch 2 Active 44mm* smartwatch during visit 1, pre-installed with the developed application (built for the Tizen mobile operating system). The application allowed participants to report mood and insulin analogue usage and automatically recorded measurements from the internal sensors (heart rate monitor, pedometer, triaxial accelerometer). First, (insulin-dependent) participants were instructed to self-report the type of insulin injected (short-acting, long-acting, or mixed insulin), dose, and time of injection through the dedicated screen. Secondly, participants were nudged four times a day at fixed times (8 AM, 12 PM, 6 PM, 8 PM) through short vibrations and activation of the smartwatch showcasing the dedicated mood screen. The screen provided the option of five moods with matching emojis: happy, relaxed, stressed, angry, and sad. Participants were instructed to respond to these nudges and to report their mood when they perceived a change in their mood. Nonetheless, participants retained the autonomy to report mood at any time via the application. An overview of the smartwatch application can be seen in Supplemental Fig. S1.

Dietary intake, both food and beverage consumption (excluding water, tea, and coffee without any additives), and physical activity were self-reported every day through the *Mijn Eetmeter* smartphone application (translates to ‘*My Eating Meter’*) on the participant’s smartphone (Voedingscentrum, The Hague, The Netherlands)^24^. Reporting dietary intake required participants to enter the meal moment (breakfast, lunch, dinner, and snacks in between these meal moments), meal contents, and amounts of these meal contents. Options for meal contents are synchronized with the Dutch Food Composition Information Portal (NVIP) from the Dutch National Institute for Public Health and the Environment^25^. The application provides predefined portion sizes per product (e.g., one standard serving of a product or the distinction between a small, medium, or large portion with an indicated weight) next to the option to report the weight of the product. Instructions were given to report mealtimes in an adjacent text field that listed the different meal moments.

Furthermore, participants were instructed to report physical activity in the application. This action consisted of selecting the type of activity from a wide range of possibilities (from walking to cleaning the house), the duration of the activity, and noting the starting time in a text field. For an impression of the application, see Supplemental Fig. S2.

### Data Security

Participants were provided with preset study *Mijn Eetmeter* accounts to promote data security and privacy, aligning with the General Data Protection Regulation (GDPR). Additionally, the smartwatch and CGM operated offline during the study period.

### Statistical Analysis

95% binomial proportion confidence intervals for wearing percentage (smartwatch was worn at the time of a nudge) and response percentage (responses to nudges when the smartwatch was determined to be worn) were calculated using Clopper-Pearson “exact” confidence intervals, unless stated otherwise. The wearing of the smartwatch was determined by the presence of a valid reading of the heart rate sensor within one minute of nudging, whereas the response percentage was defined as the percentage of responses to a nudge, within five minutes, when the smartwatch was worn according to the previous criterion.

## Data Availability

All data produced in the present study are available upon reasonable request to the authors.

## Acknowledgements

This research is part of the DiaGame project funded by NWO Data2Person (628.011.027).

## Author Contributions

R.V. was responsible for conceptualization, methodology, software, validation, formal analysis, investigation, resources, data curation, visualization, and writing the first draft of the manuscript. E.R. was responsible for conceptualization, methodology, validation, investigation, resources, and writing the first draft of the manuscript. D.C. worked on conceptualization, methodology, software, validation, investigation, resources, and data curation. E.C. was responsible for conceptualization, methodology, validation, investigation, and resources. P.G., P.W., U.K, P.H, N.R., and H.H. contributed to conceptualization, methodology, and reviewing of the manuscript. Additionally, N.R. and H.H. provided supervision and acquired funding, while N.R. was also responsible for project administration. All authors approved the definitive version of the manuscript.

## Data Availability Statement

The data used and/or analyzed in the current study are available from the corresponding author upon reasonable request.

