## Supplemental Fig. for "The DiaGame Study: Multimodal Data Collection in Free-Living Conditions Using Consumer-Level Wearable Devices and Apps in People with Diabetes"

***Supplemental Figure S1: Overview of the smartwatch application.***

*The different screens that are displayed to the participants in the smartwatch application. Participants can switch between screens by manually swiping to the left or right of the screen as shown in the figure. From left to right: screen with user information; monitoring of internal sensors; reporting insulin injections (type, dose, date, time); reporting mood with the choice of five mood categories (happy, relaxed, stressed, angry, sad); overview of previously reported insulin entries; screen with information to contact research team. Note that all information is provided in Dutch.*

*Due to the use of a non-English language (Dutch) for our study population, this figure was not allowed to be uploaded as part of the preprint. Please contact the corresponding author to request access to this information.*

***Supplemental Figure S2: Screenshots of the “Mijn Eetmeter” app demonstrating the procedures to report dietary intake and physical activity.***

*Upper panel: Steps to register the consumption of a meal or a beverage in the Mijn Eetmeter app; 1. Tap the plus icon to add a product and select the corresponding meal moment (this step is not shown, but the app will give the option of the different meal moments). 2. Search for the ingredient or product or make use of the barcode scanner. 3. Register the amount of the ingredient or product, in grams or predefined portion sizes, that was consumed. 4. Registered ingredients and products will be shown, and the time of consumption can be entered in the notes field (at the bottom) using the template inserted by the study team. Lower panel: Steps to register physical activity in the Mijn Eetmeter app; 1. Tap the plus icon to add a bout of physical activity. 2. Search for an adequate description of the activity 3. Register the duration of the activity. 4. Registered bouts of physical activity will be shown, and the starting time of the activity can be entered in the notes field (at the bottom) using the template inserted by the study team.*

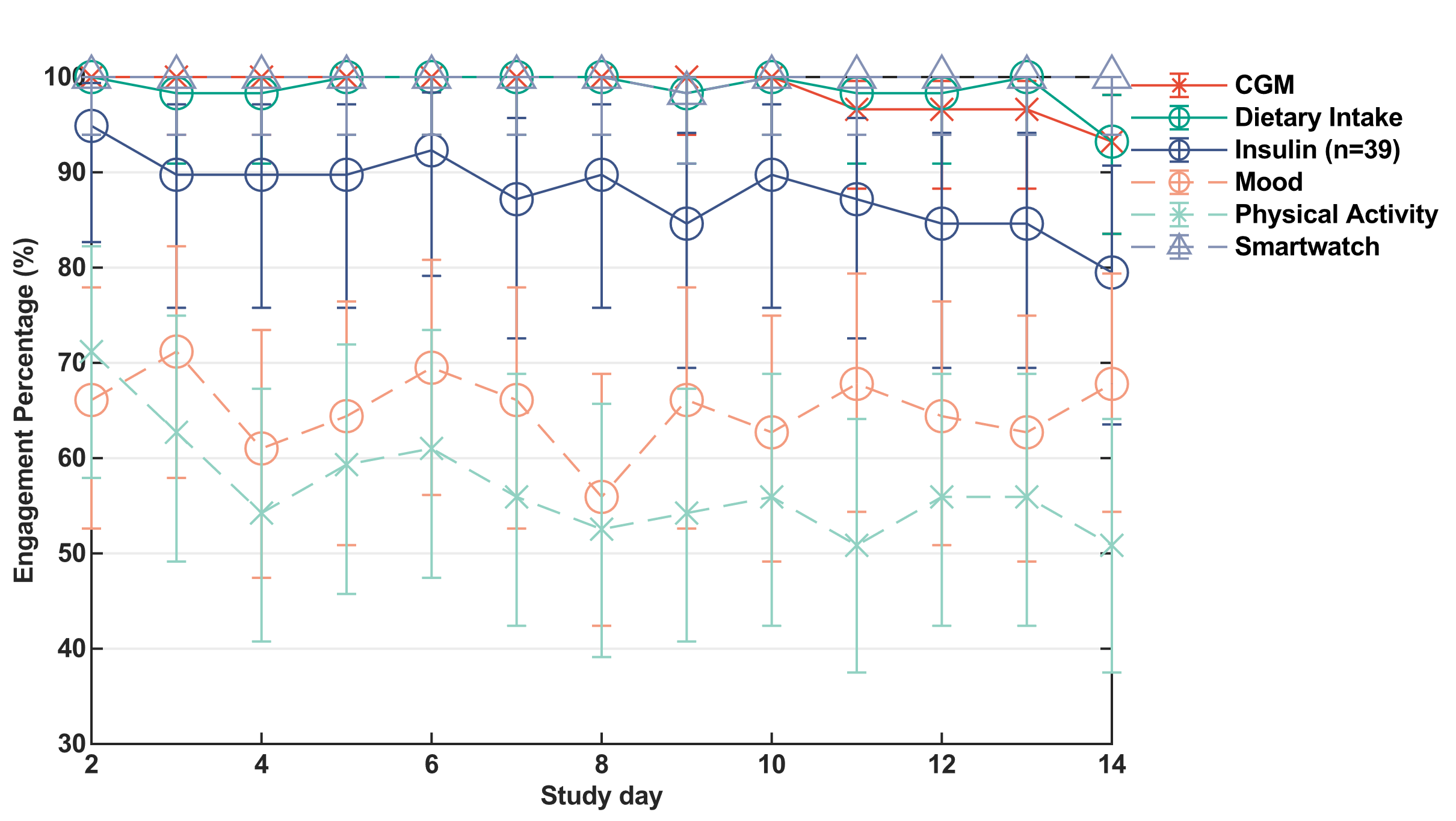

**Supplemental Figure S3: Participant engagement with devices and applications.**

Percentage of participants engaged in reporting data via the applications (dietary intake is represented by dark green circles with solid lines, insulin administration in dark blue circles with solid lines, mood in orange circles with dashed lines, physical activity in light green crosses with dashed lines) and passively collected data via the devices (CGM measurements in red crosses with solid lines, wearing of the smartwatch in light blue triangles with dashed lines) over the study period. Engagement was defined as the reporting of data on each respective variable on each study day. 95% confidence intervals are represented by the error bars. The smartwatch was considered to be worn on a day when at least 100 valid heart rate measurements were available.

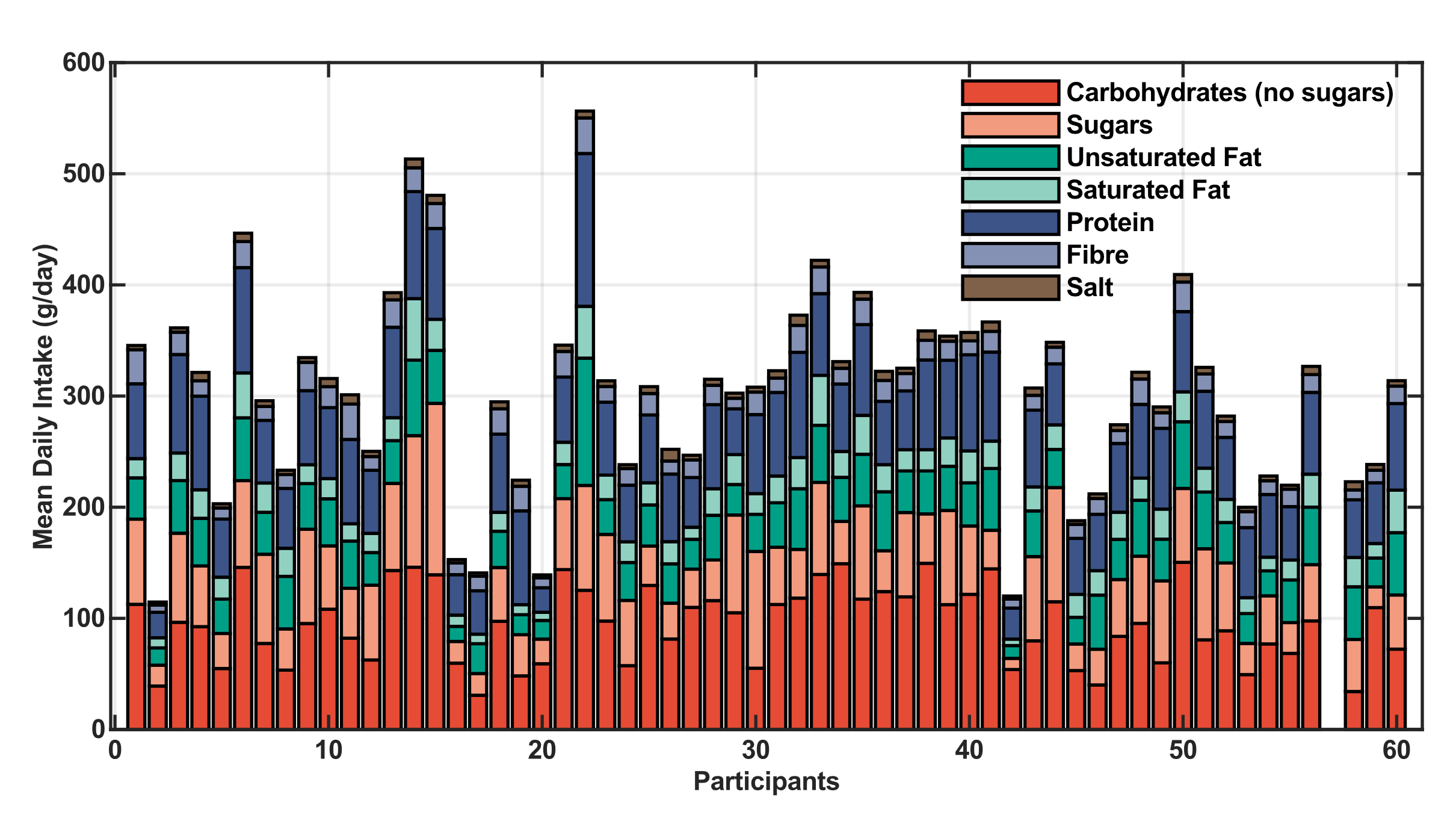

***Supplemental Figure S4: Mean daily reported macronutrient composition.***

*Mean daily reported intake of carbohydrates (excluding sugars = dark red, of which sugars = light red), fats (unsaturated fat = dark green, saturated fat = light green), protein (dark blue), fiber (light blue), and salt (dark brown) for each participant (all macronutrient intakes are in grams per day).*

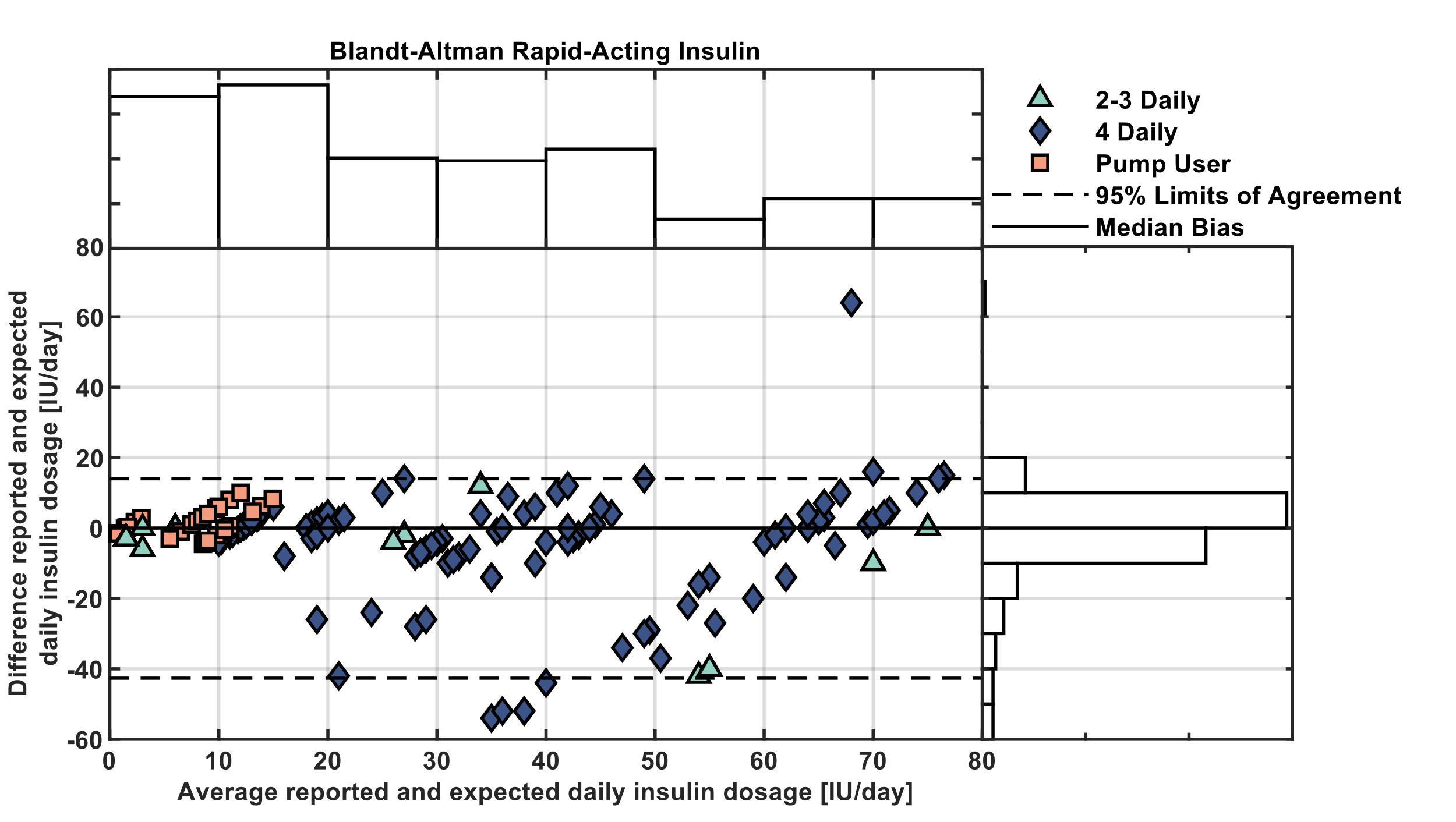

***Supplemental Figure S5: Bland-Altman plot comparing daily participant-reported rapid-acting insulin doses with expected doses from health records (n = 19).***

*Bland-Altman plot (central plot), with adjacent plots showing the marginal distributions, comparing the daily participant-reported rapid-acting insulin doses with expected doses according to their health records. Points (one per day per participant) are colored by the number of daily insulin injections for the respective participant as part of their prescribed treatment: two to three daily injections (represented by light green triangles), four injections (dark blue diamonds), and insulin pump users (light orange squares). 95% limits of agreement (dashed black lines, -43 to 14 IU/day) have been calculated using cut-off quantile estimators due to non-normal differences (Shapiro-Wilk: p < 0.05). The median bias (solid black line) was 0 IU/day. The absolute relative difference between reported and expected doses was less than 15% for 50.2% of days, whereas the relative difference was lower than -15% for 31.6% of days. Additionally, all 19 participants have at least one day with an absolute relative difference below 15%. Participants with a prescribed broad daily dose range (more than ±5 IU), who have been prescribed to inject as needed, or who did not report insulin throughout the study period have been excluded from consideration for this supplemental figure.*

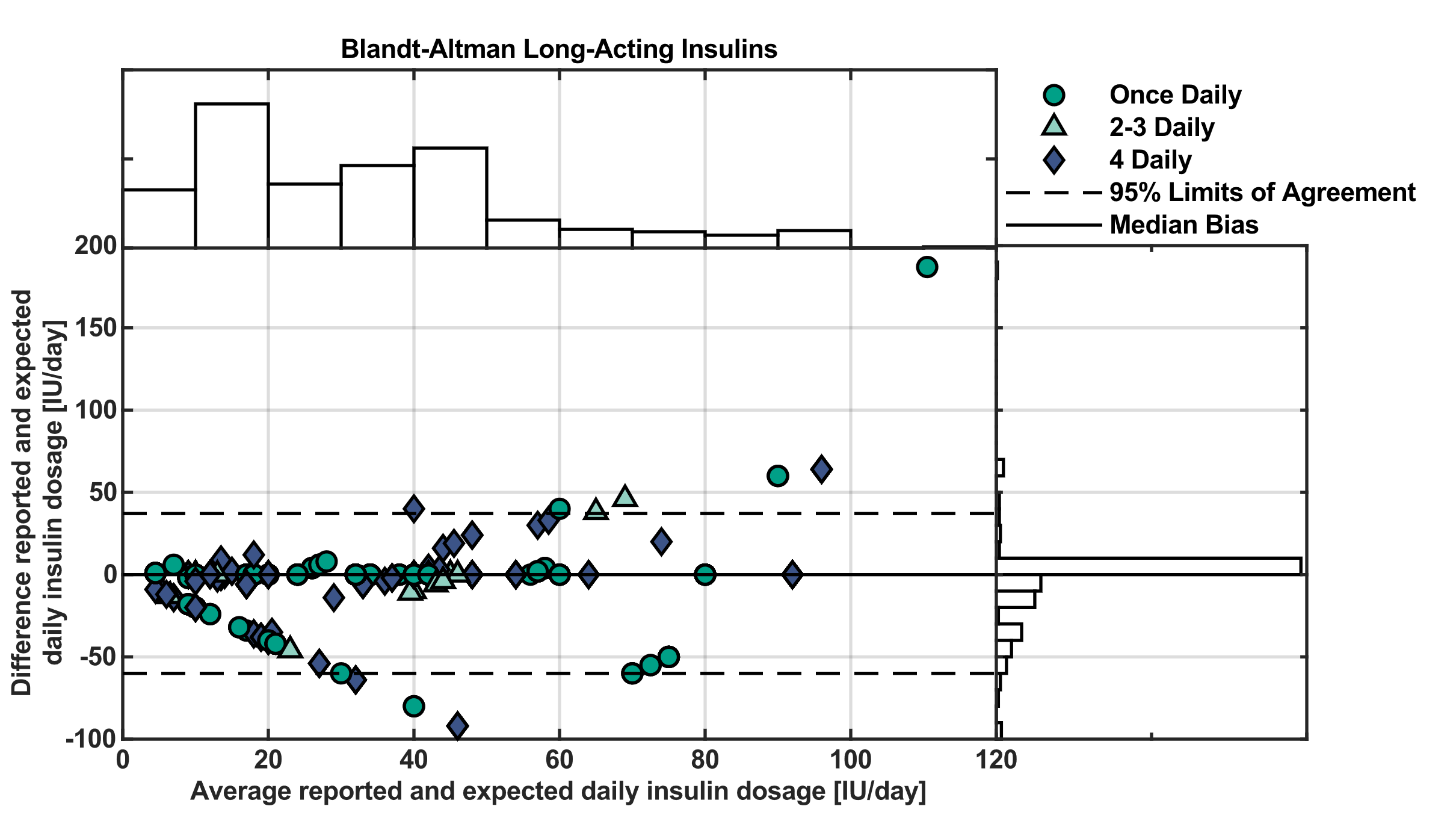

***Supplemental Figure S6: Bland-Altman plot comparing daily participant-reported ultra-long, long-acting, and intermediate insulin doses with expected doses from health records (n = 36).***

*Bland-Altman plot (central plot), with adjacent plots showing the marginal distributions, comparing the daily participant-reported ultra-long, long, and intermediate-acting insulin doses with expected doses according to their health records. Points (one per day per participant) are colored by the number of daily insulin injections for the respective participant as part of their prescribed treatment: one injection per day (dark green circles), two to three daily injections (represented by light green triangles), and four injections (dark blue diamonds). 95% limits of agreement (dashed black lines, -60 to 37 IU/day) have been calculated using cut-off quantile estimators due to non-normal differences (Shapiro-Wilk: p < 0.05). The median bias (solid black line) was 0 IU/day. The absolute relative difference between reported and expected was less than 2% for 55.5% of days, whereas the relative difference was lower than -2% for 31.0% of days. Additionally, 31 out of 36 participants have at least one day with an absolute relative difference below 2%. Participants who did not report long-acting insulin throughout the study period have been excluded from consideration for this supplemental figure.*

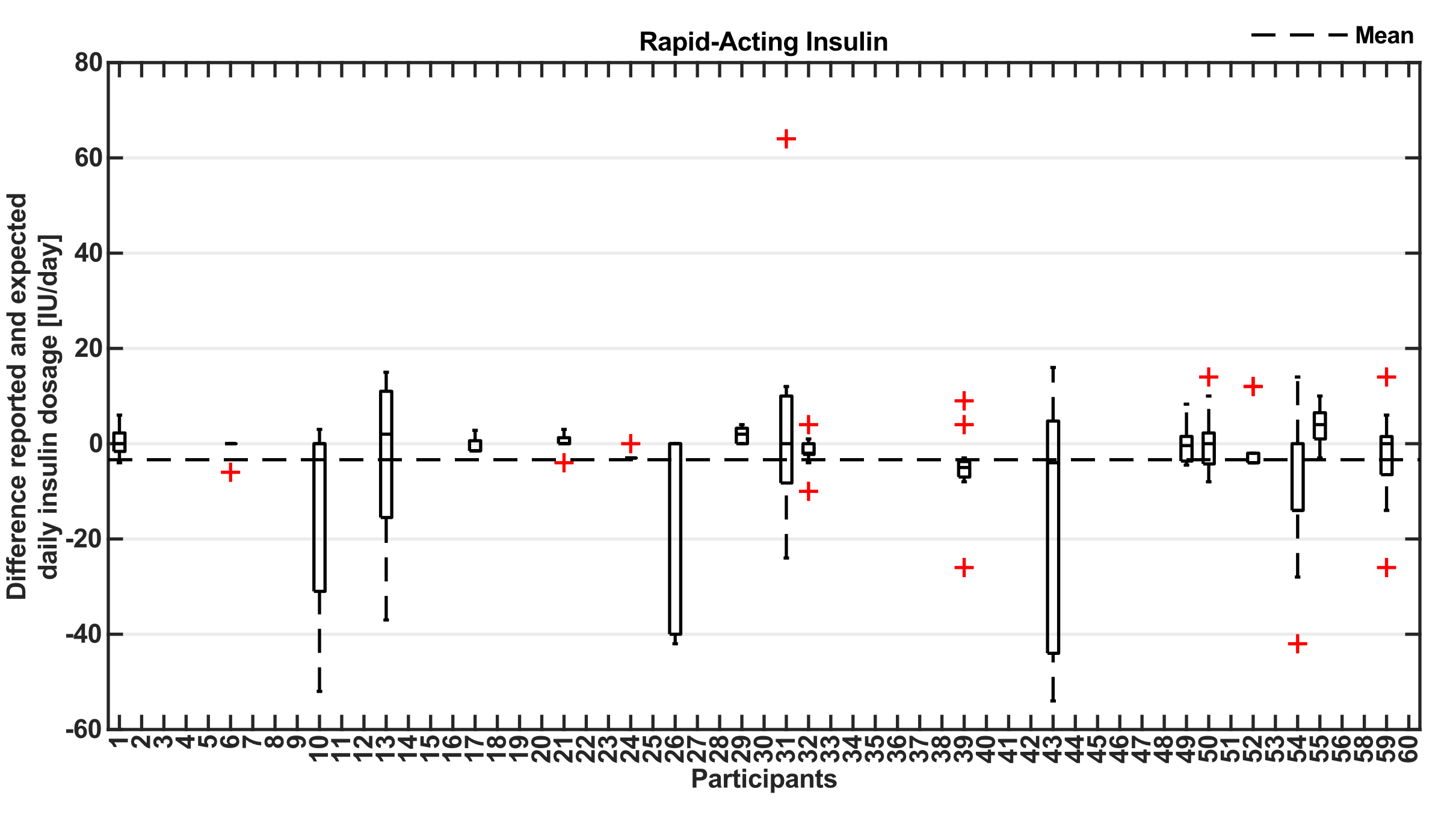

***Supplemental Figure S7: Boxplots of difference per study day between participant-reported rapid-acting insulin doses and known daily doses from health records for each participant (n = 19).***

*Boxplots of differences between daily participant-reported rapid-acting insulin doses, from study day two until study completion, and expected doses according to their health records. The overall mean difference was -3.4 IU (dashed black line, boxplot outliers are represented by red pluses). Participants with a prescribed broad dose range (more than ±5 IU), who have been prescribed to inject as needed, or who did not report insulin throughout the study period have been excluded from consideration.*

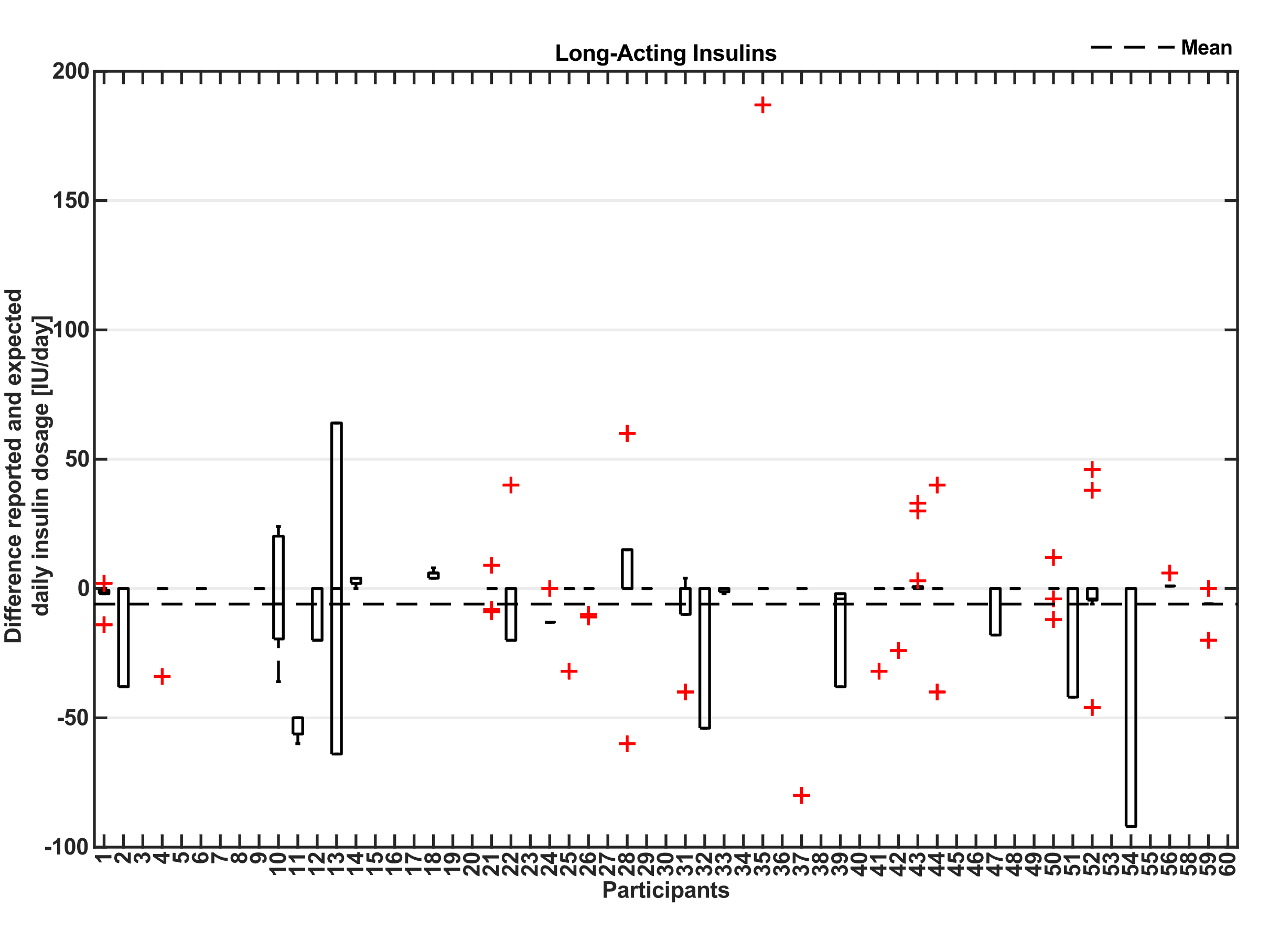

***Supplemental Figure S8: Boxplots of difference per study day between daily participant-reported ultra-long, long, and intermediate-acting insulin doses and known daily doses from health records for each participant (n = 36).***

*Boxplots of differences between daily participant-reported ultra-long, long, and intermediate-acting insulin doses, from study day two until study completion, and expected doses according to their health records. The overall mean difference was -6.0 IU (dashed black line, boxplot outliers are represented by red pluses). Participant who did not report long-acting insulin throughout the study period have been excluded from consideration.*

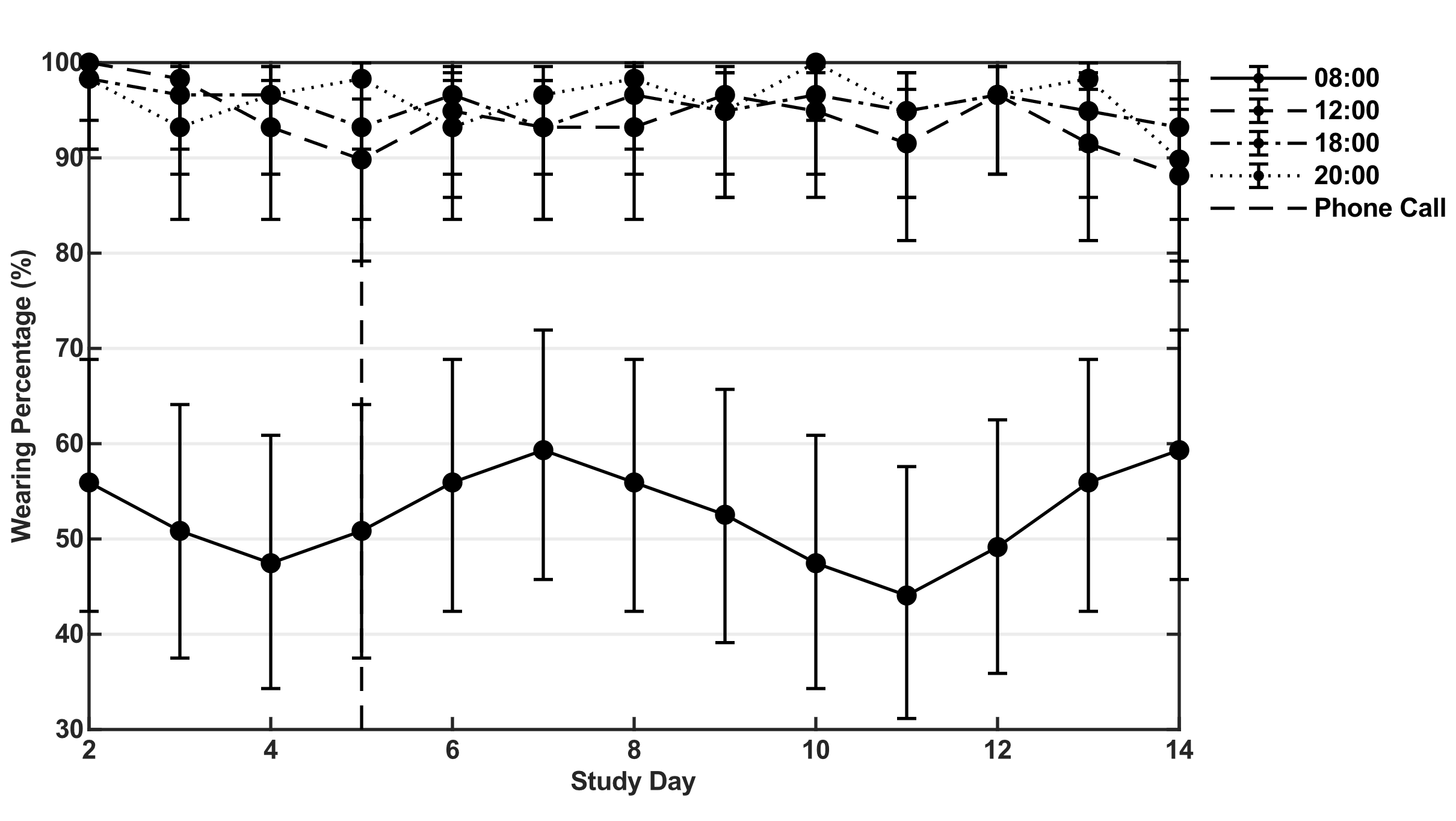

**Supplemental Figure S9: *Wearing percentage of the smartwatch over the study period at each nudge times.*** *Wearing percentage (%) of the smartwatch at each nudge time (08:00 = solid line, 12:00 = dashed line, 18:00 = dash-dot line, 20:00 = dotted line) over the study period, excluding the day of visit one, with 95% confidence intervals (error bars). Day of telephone call (can vary slightly due to the weekends) is indicated by the vertical dashed line at study day five.*

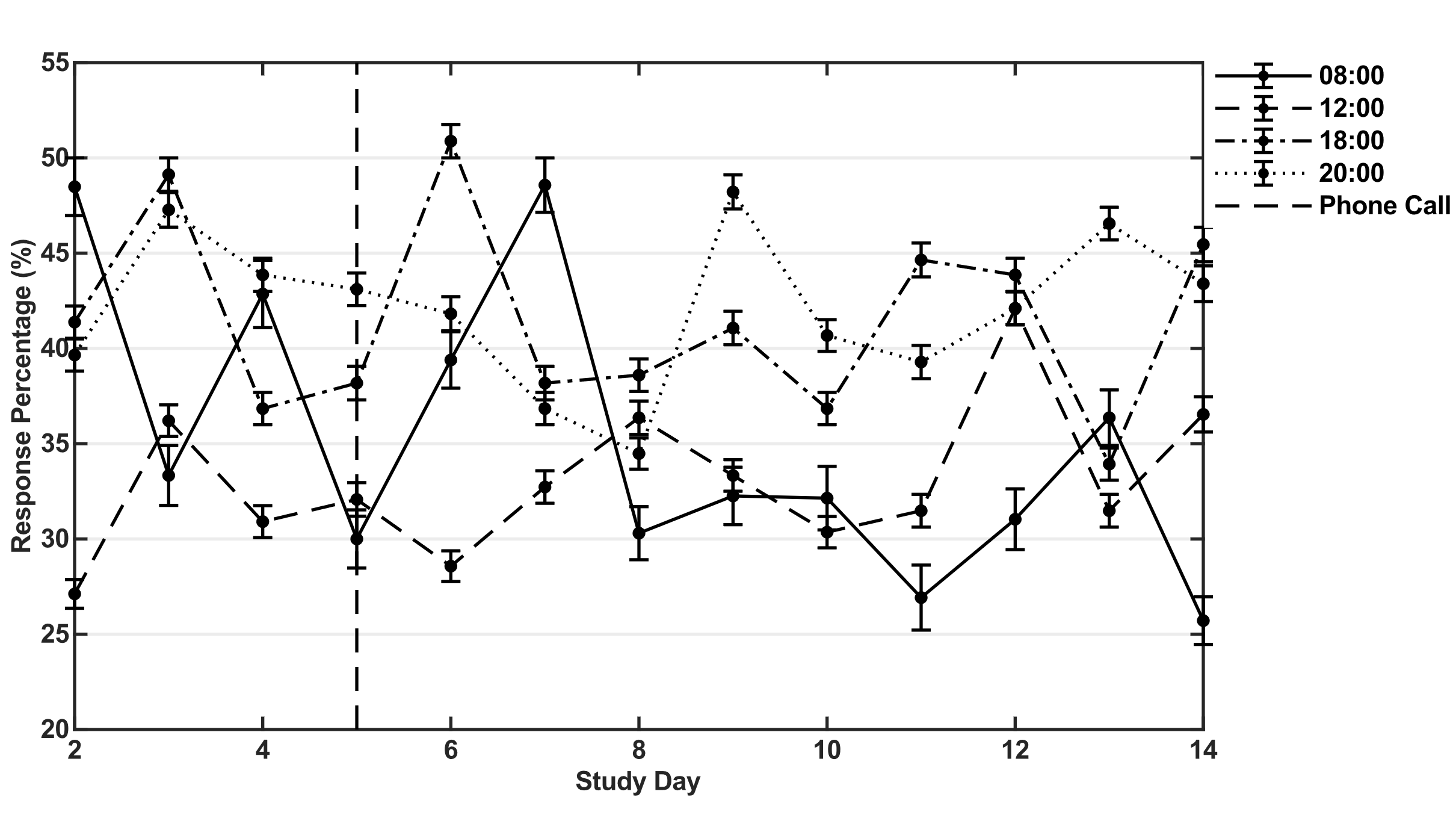

**Supplemental Figure S10: *Response percentage to the nudges over the study period at each nudge times.*** *Response percentage (%) to the nudges at each nudge time (08:00 = solid line, 12:00 = dashed line, 18:00 = dash-dot line, 20:00 = dotted line) over the study period, excluding the day of visit one, with standard error of the mean (error bars). Day of telephone call (can vary slightly due to the weekends) is indicated by the vertical dashed line at study day five.*

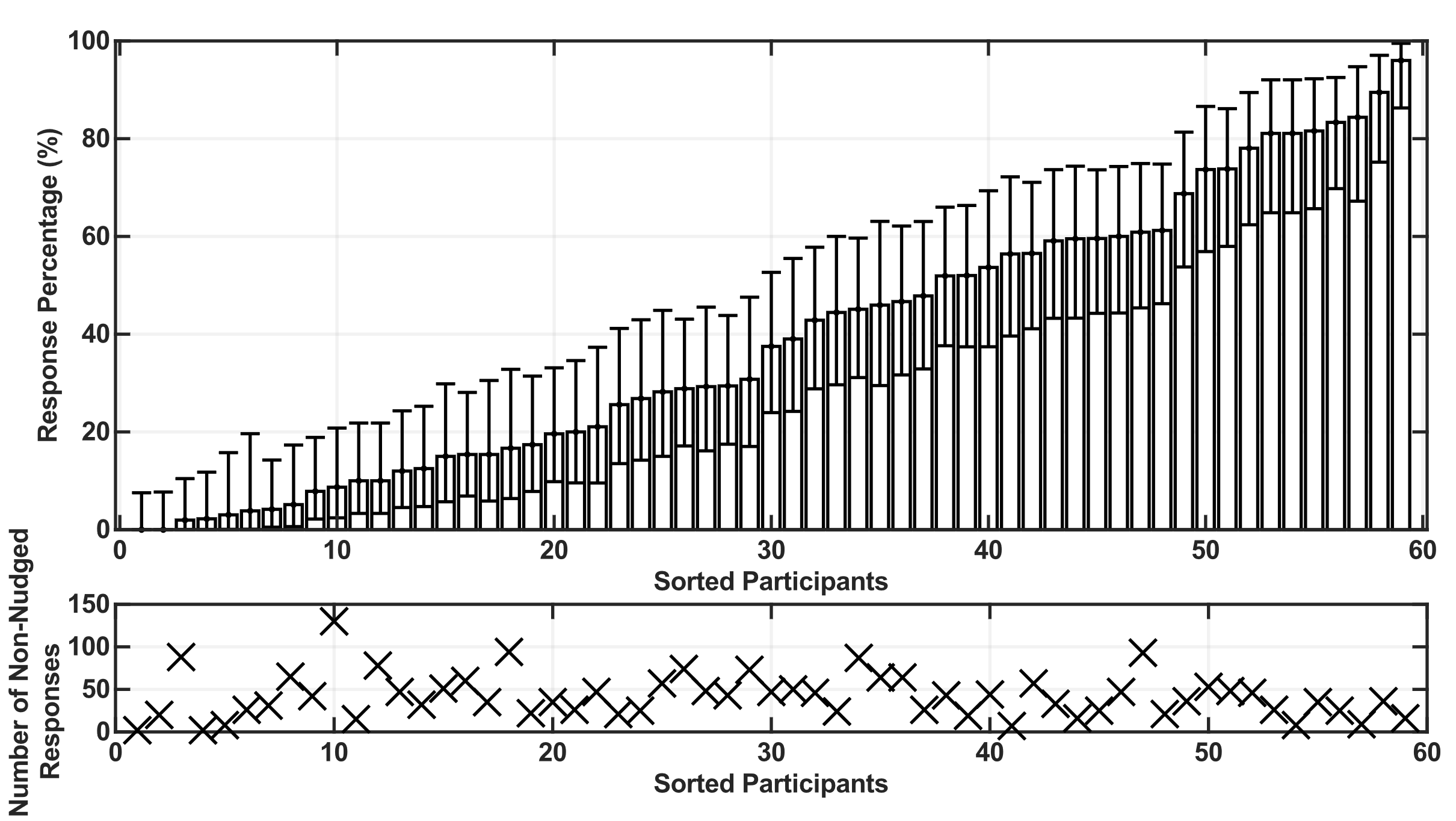

***Supplemental Figure S11:* *Overall response percentage and number of reported moods outside of the nudging periods per participant.***

*Upper panel: Sorted overall response percentage (%) per participant over the study period (bars) with 95% confidence intervals (whiskers). Lower panel: corresponding number of mood-reports outside of the nudging periods for each respective participant. The response percentages to the nudging were not correlated with the number of moods reported outside of the nudging times (Spearman’s ρ = -0.08, p = 0.53).*
